# Leveraging a Large Language Model to Assess Quality-of-Care: Monitoring ADHD Medication Side Effects

**DOI:** 10.1101/2024.04.23.24306225

**Authors:** Yair Bannett, Fatma Gunturkun, Malvika Pillai, Jessica E. Herrmann, Ingrid Luo, Lynne C. Huffman, Heidi M. Feldman

## Abstract

**Objective:** To assess the accuracy of a large language model (LLM) in measuring clinician adherence to practice guidelines for monitoring side effects after prescribing medications for children with attention-deficit/hyperactivity disorder (ADHD).

**Methods:** Retrospective population-based cohort study of electronic health records. Cohort included children aged 6-11 years with ADHD diagnosis and ≥2 ADHD medication encounters (stimulants or non-stimulants prescribed) between 2015-2022 in a community-based primary healthcare network (n=1247). To identify documentation of side effects inquiry, we trained, tested, and deployed an open-source LLM (LLaMA) on all clinical notes from ADHD-related encounters (ADHD diagnosis or ADHD medication prescription), including in-clinic/telehealth and telephone encounters (n=15,593 notes). Model performance was assessed using holdout and deployment test sets, compared to manual chart review.

**Results:** The LLaMA model achieved excellent performance in classifying notes that contain side effects inquiry (sensitivity= 87.2%, specificity=86.3/90.3%, area under curve (AUC)=0.93/0.92 on holdout/deployment test sets). Analyses revealed no model bias in relation to patient age, sex, or insurance. Mean age (SD) at first prescription was 8.8 (1.6) years; patient characteristics were similar across patients with and without documented side effects inquiry. Rates of documented side effects inquiry were lower in telephone encounters than in-clinic/telehealth encounters (51.9% vs. 73.0%, p<0.01). Side effects inquiry was documented in 61% of encounters following stimulant prescriptions and 48% of encounters following non-stimulant prescriptions (p<0.01).

**Conclusions:** Deploying an LLM on a variable set of clinical notes, including telephone notes, offered scalable measurement of quality-of-care and uncovered opportunities to improve psychopharmacological medication management in primary care.

## INTRODUCTION

Accurate measurement of clinical practice is a necessary and critical component of improving healthcare quality and health outcomes in a learning health system.^1^ However, traditional methods for capturing clinical practice, such as chart reviews, are time consuming, labor intensive, and unconducive to real-time improvement efforts.^2,3^ Large language models are a type of artificial intelligence that offers opportunities to capture clinical practice at scale by automatically analyzing free-text information from clinical notes in the electronic health records (EHR). Here, we focus on a highly prevalent childhood condition – Attention Deficit/Hyperactivity Disorder (ADHD) - which has long-standing clinical practice guidelines, as a test case for leveraging a large language model to assess quality-of-care.

ADHD is a prevalent neurodevelopmental disorder, estimated to affect 10% of US children.^4^ Most children with ADHD are treated by their primary care pediatrician (PCP).^5,6^ PCPs frequently prescribe medications, including stimulants and non-stimulants, as part of the management of ADHD. Evidence-based clinical practice guidelines for primary care management of ADHD, published and distributed by the American Academy of Pediatrics (AAP), encourage PCPs to monitor benefits and side effects when prescribing medications.^7–9^ The few studies that have assessed PCP adherence to AAP practice guidelines for ADHD medication management analyzed available EHR structured data (e.g., prescriptions, encounter dates) to assess medication prescription patterns and timely follow-up of patients.^10,11^ Studies that aimed to gather more detailed information, such as evidence in the EHR narrative text that clinicians monitored medication benefits and side effects, have been limited in scope due to the need for labor-intensive chart reviews.^11,12^

The current national quality metrics that aim to capture high-quality ADHD medication management – Healthcare Effectiveness Data and Information Set (HEDIS) measures - are claims-based metrics that are readily available for analysis and reporting at scale.^13^ However, these metrics only capture the frequency of in-office follow up of children prescribed ADHD medications. The HEDIS metrics have no evidence base (i.e., guidelines do not include a recommended frequency of follow-up) and they do not capture any guideline-based treatment recommendations, such as monitoring for medication side effects.^14^ Furthermore, a focus on information abstracted only from in-office visits may significantly underestimate the frequency of encounters. Current medication management in primary care takes place in multiple communication routes with families, including through virtual (telehealth) encounters, telephone encounters, and secure messaging systems. Indeed, one study assessing primary care medication management of ADHD found that the frequency of any type of follow-up, including communication with patients over the phone or email – not in-person follow-up alone – was associated with improved ADHD symptoms.^15^

In this study, we aimed to assess the accuracy of an open-source large language model in measuring the extent to which primary care clinicians document inquiring about ADHD medication side effects in clinical notes from in-clinic/telehealth and telephone encounters. If successful, such a model can become an integral part of robust quality measurement, highlighting avenues for near real-time improvement efforts leading to improved patient outcomes.

## METHODS

We present our study in accordance with the MINimum Information for Medical AI Reporting (MINIMAR) framework and the Strengthening the Reporting of Observational Studies in Epidemiology (STROBE) reporting guidelines.^16,17^ This study was approved by the Stanford University School of Medicine Institutional Review Board.

### Setting and population

Packard Children’s Health Alliance (PCHA) is a community-based pediatric healthcare network in the Northern California, affiliated with Stanford Children’s Health and Lucile Packard Children’s Hospital. PCHA has 25 pediatrics primary care clinics, grouped into 11 practices.

### Study design, data sources, and cohort selection

This was a retrospective population-based cohort study. We extracted structured and unstructured (free text) EHR data (2015-2022) from 11 community primary care practices of all clinic/telehealth/telephone ADHD-related encounters for patients 6-11 years. We defined an ADHD-related encounter as an encounter with an ADHD visit diagnosis or with an ADHD medication prescribed, including stimulants (methylphenidate and amphetamines) and non-stimulants (alpha-agonists and atomoxetine). ADHD diagnoses and prescriptions for ADHD medications were identified based on codes and concepts from the Observational Medical Outcomes Partnership Common Data Model (OMOP CDM), an approach that allows for common format and nomenclature across databases (see **e-supplement** for list of codes).

Since we focused on documentation of medication side effects, the study cohort included patients with ≥2 ADHD medication encounters (i.e., encounters in which the PCP prescribed stimulants or non-stimulants). The final study cohort comprised 1247 patients (see **supplementary Figure 1** for the study flowchart).

### Structured EHR data variables

We used structured data in the EHR to describe the following patient characteristics: patient age (at encounter of interest), sex, race/ethnicity (Asian/Non-Hispanic, Black/Non-Hispanic, Hispanic, White/Non-Hispanic, Other/Non-Hispanic, Unknown), and medical insurance at first ADHD medication encounter (private/public). In our analyses, we separated patients into two age groups (6-7 or 8-11 years at the time of the first prescription) based on our clinical experience that patients under 8 years have a higher rate of ADHD medication side effects than those who are 8 years and older.

### Manual chart review and annotation of clinical notes

The primary outcome of interest was the rate of documentation inquiring about medication side effects in clinical notes of ADHD-related encounters occurring within 3 months of an ADHD medication prescription, starting from the second medication encounter onwards per patient, when monitoring of medication side effects is expected. Side effects inquiry included documentation of either absence of side effects (e.g., “no weight loss”) or presence of side effects (e.g., “reduced appetite”). We included clinical notes from all in-clinic, telehealth, and telephone ADHD-related encounters conducted after an ADHD medication was first prescribed (4369 in-clinic/telehealth notes and 11,224 telephone notes; total notes n=15,593).

To create a “ground truth” for documentation of side effects inquiry, we developed annotation guidelines used by two clinicians (YB, JH) who performed independent chart review and annotation of a sample of clinical notes using the CLAMP software.^18^ We sampled and annotated 2-26 clinical notes per patient from medication encounters (including in-clinic/telehealth and telephone encounters) for a sample of 119 patients (n=501 notes). Inter-annotator agreement (IAA) was assessed using the Cohen’s kappa statistic.^19^ After confirming IAA=0.86 for the first 84 notes, the annotators divided the annotation of the remaining notes (n=417).

### Large language model development and train/test split

We developed a binary classification pipeline based on the open-source Large Language Model Meta AI, trained on 13 billion parameters (LLaMA13B), to classify notes as containing or not containing documentation of side effects inquiry (see **e-methods supplement** describing model architecture and optimization). **Figure 1** illustrates the model training and deployment workflow. The annotated set of 501 notes was subdivided with an 80-20 split into train (n=411) and holdout test (n=90) sets. The train set was used for model development and hyperparameter tuning while the holdout test set was set aside to evaluate model performance compared to ground truth labels using various metrics including sensitivity, specificity, and area under the receiver operating characteristic curve (AUC). Model thresholds were selected to maximize sensitivity and minimize the false negative rate on the training set because we wanted to avoid wrongly classifying notes to suggest clinicians as not adhering to practice guidelines when they actually did. We report 95% confidence intervals (CI) for AUC, which captures the discrimination power of the model and is not influenced by threshold selection. After confirming acceptable model performance on the holdout test set, we deployed the model on the remaining unannotated notes from all ADHD-related encounters for the study cohort (deployment test set, n=15,184). We then sampled and annotated 366 notes (IAA=0.93) to assess model performance in the deployment test set. The code is available via Github at https://github.com/ybannett/NLP_ADHD_SEI.

**Figure 1.**
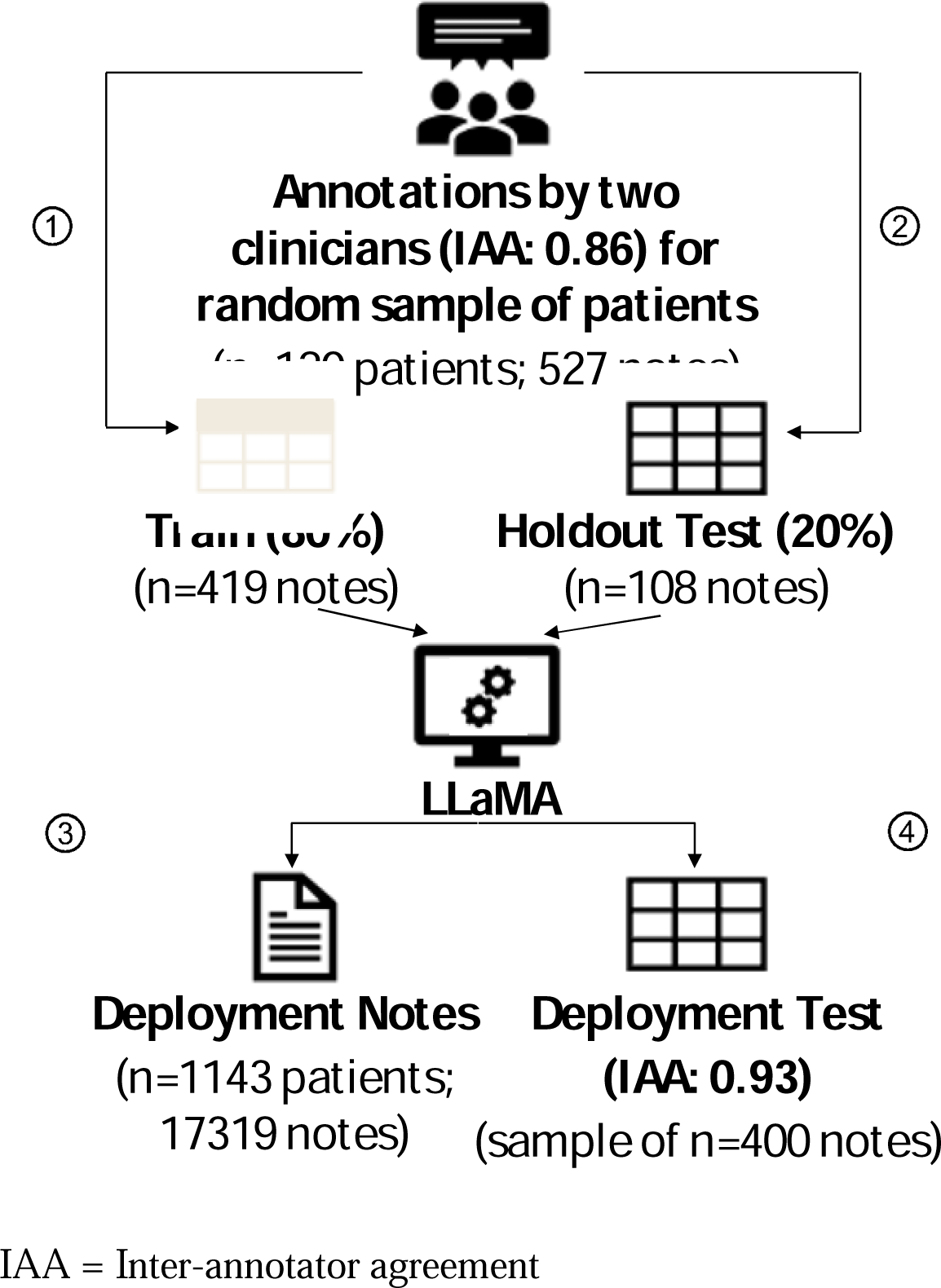
Model training, testing, and deployment.

### Error and fairness analysis

For the error analysis, the misclassified notes in the holdout and deployment test sets were reviewed to understand model errors and potential reasoning behind misclassifications. For the fairness analysis, we utilized the classification parity approach to assess whether model outcomes are roughly equal across several patient subgroups divided by patient attributes that included insurance type, sex, and age at first prescription.^20^ The fairness analysis was completed to rule out model bias that may perpetuate disparities in care, if such disparities exist.^21,22^ We did not examine race/ethnicity data in the fairness analysis due to a large percentage of missing data and co-linearity between insurance type and race/ethnicity in our data.^23^

### Statistical analysis

Continuous variables were summarized by the mean and standard deviation, and categorical variables were presented as counts and percentages. The balance of demographic characteristics between the groups was assessed using the absolute standardized differences (ASD). ASD values of 0.2, 0.5, and 0.8 correspond to small, medium, and large differences between the groups, respectively. A two-sided proportion test with a significance level of 0.05 was used to compare the proportion of encounters with side effect inquiries between 1) telephone encounters and in-clinic/telehealth encounters, and 2) encounters associated with stimulant prescriptions, non-stimulant prescriptions, or both. Only race/ethnicity had missing data (29.5%). All analyses were conducted using Python version 3.11.5.

## RESULTS

### Model performance

The LLaMA model achieved excellent performance in classifying notes that contain side effects inquiry, as compared to ground truth labels. In the holdout test set (n=90 notes), the model achieved sensitivity of 87.2%, specificity of 86.3%, and an AUC of 0.93 (CI^95%^: 0.88-0.99). In the deployment test set (n=366 notes), the model achieved sensitivity of 87.2%, specificity of 90.3%, and an AUC of 0.92 (CI^95%^: 0.89-0.94).

### Error analysis

In an error analysis, we investigated potential reasons for model misclassifications. False positive classifications included documentations that were not clearly related to the prescribed ADHD medication (e.g., review of systems) or documented side effects from other prescribed medications (e.g., medication for acne). False negative classifications included abbreviated documentation (e.g., “follow up add meds weight loss”) or documentation of non-specific symptoms (e.g., “giggly and disruptive on new med”).

### Fairness analysis

As illustrated in **Figure 2**, the model exhibited similar AUC values (presented with 95% CI) across different patient subgroups, including patient age group, sex, and insurance type.

**Figure 2.**
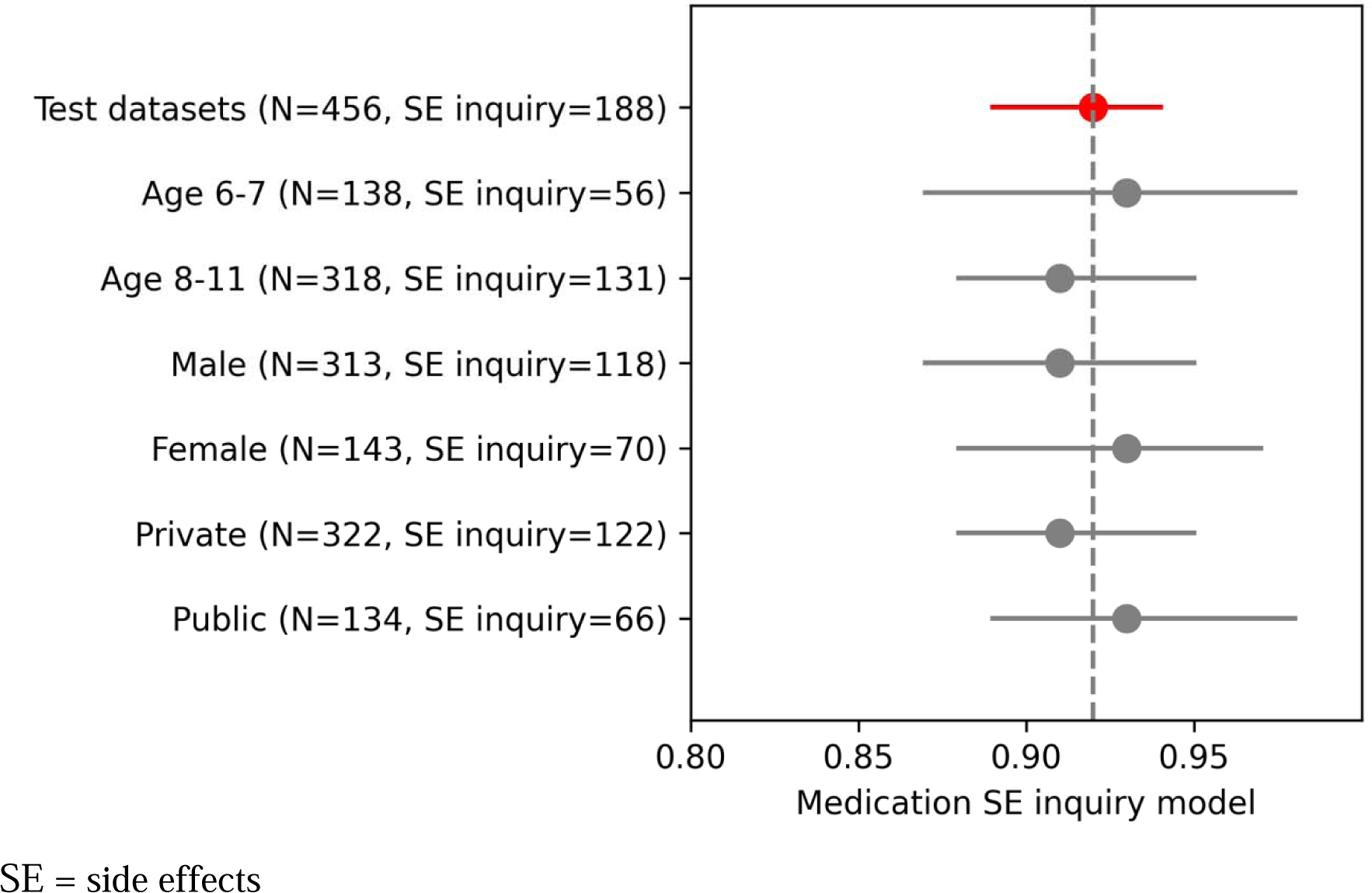
Fairness analysis results.

### Model findings and study cohort

The study cohort included 1247 patients aged 6-11 years who had at least two primary care visits in which an ADHD medication was prescribed by the PCP. Cohort characteristics by documentation of side effects inquiry (yes/no), based on model classifications, are displayed in **Table 1**. Of 1247 patients, 72.6% (n=917) were first prescribed medications when they were 8-11 years of age and 73.9% (n=922) were male. The cohort primarily consisted of White/Non-Hispanic patients (n=549, 44.0%) or Unknown race/ethnicity patients (n=365, 29.3%). Most patients were privately insured (n=929, 74.5%). Based on model classification, in 85.6% of patients (n=1067) the PCP documented inquiring about medication side effects in at least one ADHD-related encounter (starting at the second medication encounter per patient). Rates of documented side effect inquiries were higher for patients with public insurance compared to those with private insurance (90.6% vs 83.9%, absolute standardized difference=0.24), while no differences were observed with regard to age group, sex, or race/ethnicity.

**Table 1.** Characteristics of study cohort: Patients ages 6-11 years prescribed medications for ADHD by a primary care pediatrician.

|  | Overall<br>N=1247 | No Side Effects<br>Inquiry<br>N=180 | Side Effects<br>Inquiry<sup>b</sup><br>N=1067 | Absolute<br>Standardized<br>Difference<sup>c</sup> |
| --- | --- | --- | --- | --- |
| **Age at diagnosis, N (%)<sup>a</sup>** |  |  |  |  |
| 6-7 years | 342 (27.4) | 37 (20.6) | 305 (28.6) | 0.18 |
| 8-11 years | 905 (72.6) | 143 (79.4) | 762 (71.4) |  |
| **Age at 1st prescription, N (%)** |  |  |  |  |
| 6-7 years | 293 (23.5) | 30 (16.7) | 263 (24.6) | 0.19 |
| 8-11 years | 954 (76.5) | 150 (83.3) | 804 (75.4) |  |
| **Sex (Male), N (%)** | 922 (73.9) | 131 (73.6) | 791 (74.1) | 0.01 |
| **Race/Ethnicity, N (%)** |  |  |  |  |
| Asian /Non-Hispanic | 74 (5.9) | 9 (5.0) | 65 (6.1) | 0.05 |
| Black /Non-Hispanic | 60 (4.8) | 3 (1.7) | 57 (5.3) | 0.17 |
| Hispanic | 190 (15.2) | 30 (16.7) | 160 (15.0) | 0.05 |
| Other /Non-Hispanic | 9 (0.7) | 0 (0.0) | 9 (0.8) | 0.10 |
| White /Non-Hispanic | 549 (44.0) | 71 (39.4) | 478 (44.8) | 0.11 |
| Unknown | 365 (29.3) | 67 (37.2) | 298 (27.9) | 0.20 |
| **Insurance, N (%)** |  |  |  |  |
| Private | 929 (74.5) | 150 (83.3) | 779 (73.0) | 0.24 |
| Public | 318 (25.5) | 30 (16.7) | 288 (27.0) |  |

<sup>a</sup>Age at first encounter with an ADHD diagnosis within the patient age range.
<sup>b</sup>Documented inquiry of side effects, as classified by the LLaMA model, in at least one ADHD-related encounter starting at the second time an ADHD medication was prescribed.
<sup>c</sup>Absolute Standardized Difference values of 0.2, 0.5, and 0.8 correspond to small, moderate, and large differences, respectively.

### Model findings by primary care practice, encounter modality, and medication type

Overall, side effects inquiry was documented in 59.8% (n=5884) of ADHD-related encounters and was highly variable across primary care practices (range 14-74% of encounters, **Figure 3**). More than 50% of ADHD-related encounters were completed by telephone in 7 out of 11 practices. However, the proportion of telephone encounters with documented side effects inquiry was significantly lower at 52.0% compared to 72.9% for in-clinic/telehealth encounters (p<0.01), with high inquiry rates by telephone observed in only two practices. When examining encounters by the type of ADHD medication prescribed, side effects inquiry was documented in 61.1% (n= 5440) of ADHD-related encounters within three months after stimulants were prescribed and in 48.2% (n=197) of encounters after non-stimulants were prescribed (p<0.01, **Figure 4**). Side effects inquiry was documented in 46.8% (n=247) of encounters after both stimulants and non-stimulants were prescribed.

**Figure 3.**
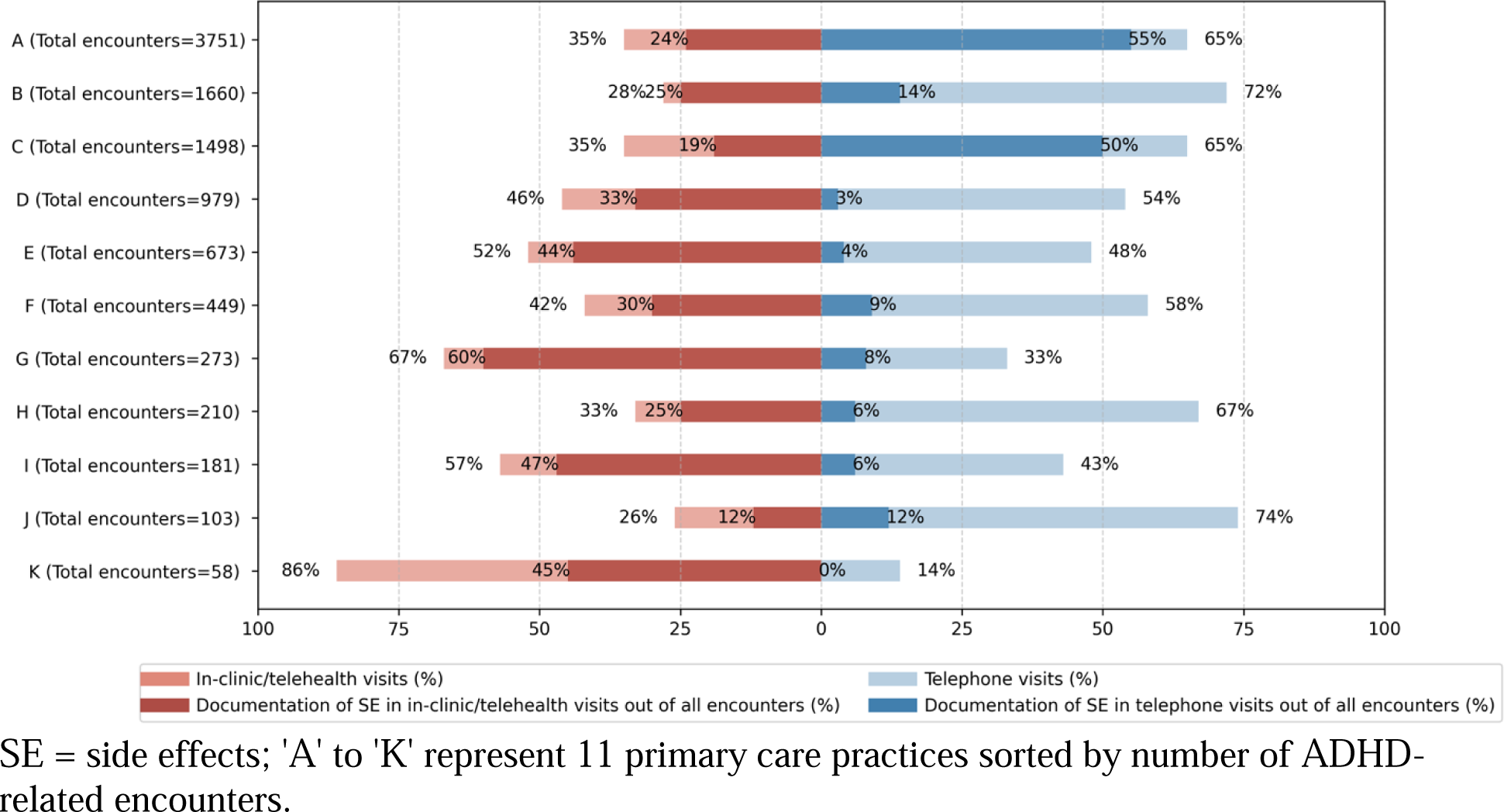
ADHD-related encounters with and without side effects (SE) inquiry by primary care practice.

**Figure 4.**
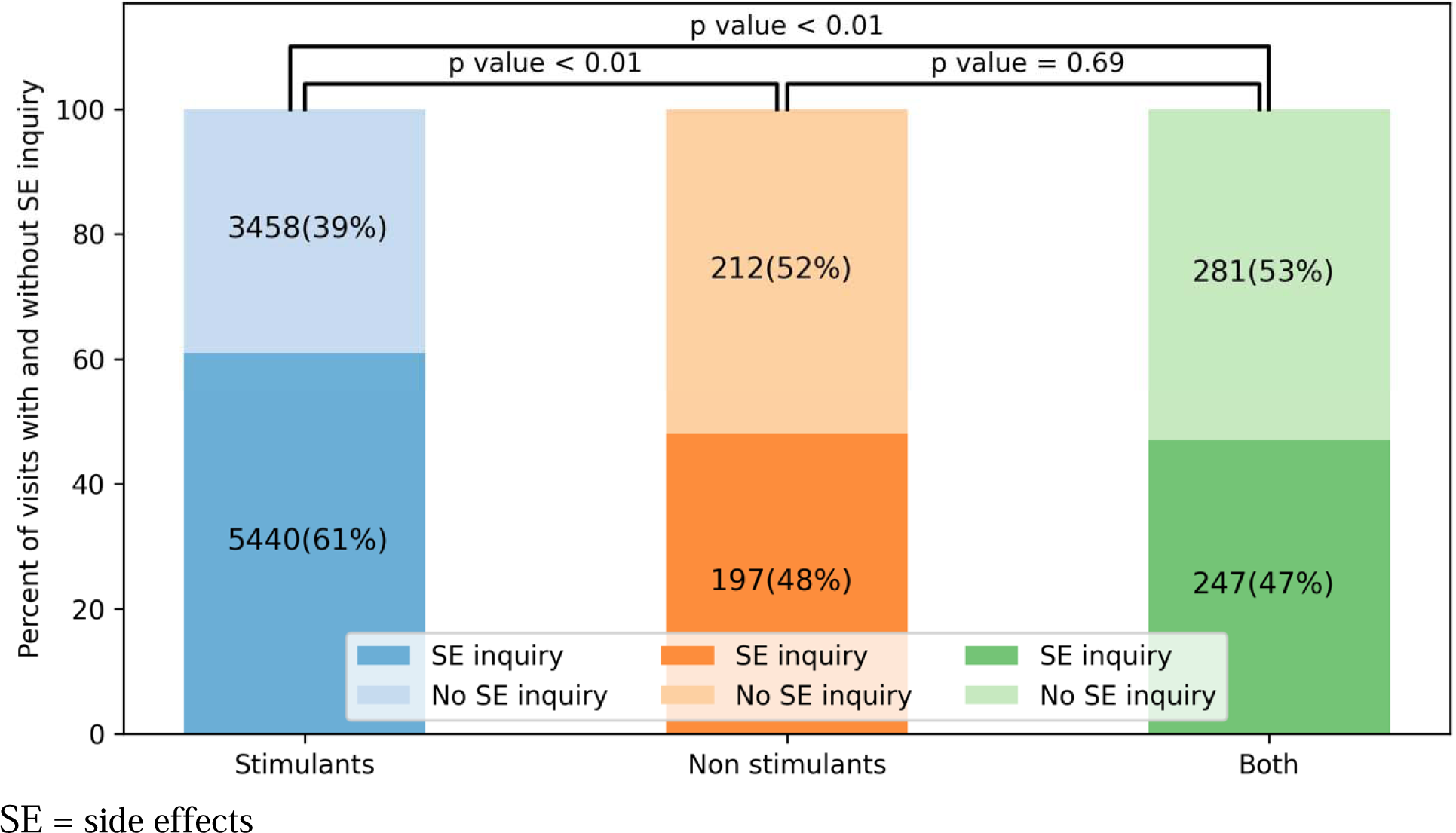
ADHD-related encounters with and without side effects (SE) inquiry by medication type.

## DISCUSSION

In this study, we have shown that deploying a large language model (LLM) on different types of clinical notes can effectively assess adherence to AAP practice guidelines for medication management of children with ADHD, a highly prevalent condition. Fairness analysis did not indicate any evidence of model bias. By deploying this model, we uncovered specific targets for improvement in PCP medication management, including under-utilization of telephone encounters to monitor medication side effects, and low rates of side effects management when prescribing non-stimulants.

The high LLM performance rates established in this study demonstrate that LLMs offer an accurate and reliable method of analyzing the vast amount of unstructured EHR data that represents the documentation trail of current pediatric care, which includes significant between-visit management of chronic conditions (e.g., telephone encounters, secure messaging). Our findings are promising in that LLMs allow efficient data mining and interpretation of large volumes of clinical data that, thus far, have been inaccessible through existing chart extraction methodologies. Additionally, the automaticity and speed of LLMs offers the opportunity to develop near real-time feedback through dashboards for clinicians and health organizations on their current care – an effective method to improve clinical practice.^24^

When applying a large language model to assess documentation of clinical care, it is critical to assess AI model bias given its potential to perpetuate health disparities.^21,22^ We therefore incorporated a fairness analysis into our study. In this case, we confirmed that the model performance did not differ significantly across patient subgroups, including patient age group, sex, and insurance type.

Several targets for quality improvement were uncovered by deploying the LLM on all clinical notes. While all practices had a high utilization of telephone visits for ADHD medication management, only two of the 11 practices used telephone visits to regularly inquire about medication side effects. This finding serves as a learning opportunity for other practices. A simple intervention that adopts a standardized medication refill form, currently used in the two practices that documented side effects inquiry in most telephone encounters, can have a significant positive effect on the quality of medication management. Another finding was the lower rates of side-effects inquiry in children prescribed non-stimulants, as compared to stimulants. Here too, an intervention that targets potential knowledge gaps related to side effects profile of these less frequently prescribed medications can promote high quality care.

This study complements our previous study that focused on clinician adherence to AAP guidelines in recommending non-pharmacological behavioral treatment for young children with ADHD, in which we demonstrated the use of large language models in the evaluation of quality-of-care for children with ADHD.^25^ These two studies are the first, to our knowledge, that provide objective support for the successful use of artificial intelligence (AI) to provide a comprehensive evaluation of ADHD management, a prevalent neurobehavioral condition that is predominantly managed in primary care. Future implementation of such algorithms, could allow primary care clinicians to receive feedback on between-visit management (e.g., telephone encounters for medication management) and to receive real-time decision support (e.g., prompts for inquiring about specific side effects) – two areas of need in primary care that have been recently identified as areas that could benefit from AI.^26^

### Limitations

Our cohort identification relied on multiple medication prescriptions with a primary indication for ADHD. Although this approach facilitates standardized implementation in any healthcare system, it introduces some level of misclassification error. Our assessment of quality-of-care in this study relies on clinician documentation in the EHR. It is possible that, in some cases, clinicians inquired about side effects without documenting their inquiries in the EHR.

Furthermore, our study focused on medication management of ADHD by PCPs, and we did not have information on prescriptions provided to children outside of the examined network (e.g., by child psychiatrists). Because we were interested in measuring clinician adherence to guidelines, our outcome included any mention of side effects inquiry; we did not have information on rates of side effects presence and absence. We are currently examining the use of another model to answer this separate research question and use case. Finally, although the study was conducted in a large network of primary care practices with a diverse population (including 15% Hispanic, similar to the US census), its generalizability and the accuracy of model classifications needs to be assessed in other healthcare networks.

## CONCLUSION

Given the high prevalence of ADHD and the potential harm that can be caused to children and adolescents if psychopharmacological medication side effects are not considered and documented as part of clinical care, this study carries significant implications. By leveraging recent advances in the ability of large language models to accurately classify large bodies of text, our novel approach offers an efficient way to assess the quality of ADHD medication management. The rapid transformation of clinical data into knowledge can provide clinicians and healthcare organizations with timely and actionable feedback. We are currently working to expand this approach to include patient vitals data (i.e., weight, blood pressure, pulse) and documentation of patient care through the EHR secure messaging system, which were not available in the dataset created for this study, representing an additional step towards a comprehensive assessment of care for a large population of patients. Following replication of this approach in other healthcare systems, implementation of LLMs can contribute to enhancing evidence-based and equitable healthcare delivery for children with ADHD, ultimately improving patient outcomes.

## Supporting information

eSupplement

## Data Availability

The entire code for the pipeline and training of the large language model, which can be used to reproduce our study in other settings, is available in the GitHub repository at https://github.com/ybannett/NLP_ADHD_SEI. The datasets generated and analyzed in the current study contain protected patient health information and are therefore not publicly available; the data will be shared on reasonable request to the corresponding author.

## Acknowledgments

This research used data and services provided by STARR, “STAnford medicine Research data Repository,” a clinical data warehouse containing live Epic data from Stanford Health Care (SHC), the Stanford Children’s Hospital (SCH), the University Healthcare Alliance (UHA) and Packard Children’s Health Alliance (PCHA) clinics and other auxiliary data from Hospital applications such as radiology PACS. STARR platform is developed and operated by Stanford Medicine Research IT team and is made possible by Stanford School of Medicine Research Office. We thank Packard Children’s Health Alliance and Stanford Research Information Technology for their support and assistance in data acquisition and extraction.

## References

1. Etheredge LM. A rapid-learning health system. Health Aff (Millwood). Mar-Apr 2007;26(2):w107-18. doi:10.1377/hlthaff.26.2.w107

2. Zima BT, Mangione-Smith R. Gaps in quality measures for child mental health care: an opportunity for a collaborative agenda. Journal of the American Academy of Child and Adolescent Psychiatry. Aug 2011;50(8):735–7. doi:10.1016/j.jaac.2011.05.006

3. Schuster MA, Onorato SE, Meltzer DO. Measuring the Cost of Quality Measurement: A Missing Link in Quality Strategy. Jama. Oct 3 2017;318(13):1219–1220. doi:10.1001/jama.2017.11525

4. Li Y, Yan X, Li Q, et al. Prevalence and Trends in Diagnosed ADHD Among US Children and Adolescents, 2017-2022. JAMA Netw Open. Oct 2 2023;6(10):e2336872. doi:10.1001/jamanetworkopen.2023.36872

5. Visser SN, Bitsko RH, Danielson ML, et al. Treatment of Attention Deficit/Hyperactivity Disorder among Children with Special Health Care Needs. J Pediatr. Jun 2015;166(6):1423–30.e1-2. doi:10.1016/j.jpeds.2015.02.018

6. Albert M, Rui P, Ashman JJ. Physician Office Visits for Attention-deficit/Hyperactivity Disorder in Children and Adolescents Aged 4–17 Years: United States, 2012-2013. NCHS data brief. Jan 2017;(269):1-8.

7. Clinical practice guideline: treatment of the school-aged child with attention-deficit/hyperactivity disorder. Pediatrics. Oct 2001;108(4):1033-44.

8. Wolraich M, Brown L, Brown RT, et al. ADHD: clinical practice guideline for the diagnosis, evaluation, and treatment of attention-deficit/hyperactivity disorder in children and adolescents. Pediatrics. Nov 2011;128(5):1007–22. doi:10.1542/peds.2011-2654

9. Wolraich ML, Hagan JF, Jr., Allan C, et al. Clinical Practice Guideline for the Diagnosis, Evaluation, and Treatment of Attention-Deficit/Hyperactivity Disorder in Children and Adolescents. Pediatrics. Oct 2019;144(4)doi:10.1542/peds.2019-2528

10. Bannett Y, Feldman HM, Gardner RM, et al. Attention-Deficit/Hyperactivity Disorder in 2-to 5-Year-Olds: A Primary Care Network Experience. Acad Pediatr. Apr 28 2020;doi:10.1016/j.acap.2020.04.009

11. Epstein JN, Kelleher KJ, Baum R, et al. Variability in ADHD care in community-based pediatrics. Pediatrics. Dec 2014;134(6):1136–43. doi:10.1542/peds.2014-1500

12. Gordon MK, Baum RA, Gardner W, et al. Comparison of Performance on ADHD Quality of Care Indicators: Practitioner Self-Report Versus Chart Review. Journal of attention disorders. Aug 2020;24(10):1457–1461. doi:10.1177/1087054715624227

13. National Committee for Quality Assurance. Follow-up care for children prescribed ADHD medication. Accessed October 24, 2019, Available at: http://www.ncqa.org.

14. Zima BT, Murphy JM, Scholle SH, et al. National quality measures for child mental health care: background, progress, and next steps. Pediatrics. Mar 2013;131 Suppl 1:S38–49. doi:10.1542/peds.2012-1427e

15. Epstein JN, Kelleher KJ, Baum R, et al. Specific Components of Pediatricians’ Medication-Related Care Predict Attention-Deficit/Hyperactivity Disorder Symptom Improvement. Journal of the American Academy of Child and Adolescent Psychiatry. Jun 2017;56(6):483–490.e1. doi:10.1016/j.jaac.2017.03.014

16. Hernandez-Boussard T, Bozkurt S, Ioannidis JPA, et al. MINIMAR (MINimum Information for Medical AI Reporting): Developing reporting standards for artificial intelligence in health care. Journal of the American Medical Informatics Association: JAMIA. Dec 9 2020;27(12):2011–2015. doi:10.1093/jamia/ocaa088

17. Vandenbroucke JP, von Elm E, Altman DG, et al. Strengthening the Reporting of Observational Studies in Epidemiology (STROBE): explanation and elaboration. PLoS Med. Oct 16 2007;4(10):e297. doi:10.1371/journal.pmed.0040297

18. Soysal E, Wang J, Jiang M, et al. CLAMP – a toolkit for efficiently building customized clinical natural language processing pipelines. Journal of the American Medical Informatics Association. 2017;25(3):331–336. doi:10.1093/jamia/ocx132

19. McHugh ML. Interrater reliability: the kappa statistic. Biochemia medica. 2012;22(3):276–282.

20. Röösli E, Bozkurt S, Hernandez-Boussard T. Peeking into a black box, the fairness and generalizability of a MIMIC-III benchmarking model. Sci Data. Jan 24 2022;9(1):24. doi:10.1038/s41597-021-01110-7

21. Czarnowska P, Vyas Y, Shah K. Quantifying Social Biases in NLP: A Generalization and Empirical Comparison of Extrinsic Fairness Metrics. Transactions of the Association for Computational Linguistics. 2021;9:1249–1267. doi:10.1162/tacl_a_00425

22. Röösli E, Rice B, Hernandez-Boussard T. Bias at warp speed: how AI may contribute to the disparities gap in the time of COVID-19. Journal of the American Medical Informatics Association. 2020;28(1):190–192. doi:10.1093/jamia/ocaa210

23. Bannett Y, Gardner RM, Huffman LC, et al. Continuity of Care in Primary Care for Young Children With Chronic Conditions. Academic Pediatrics. 2023;23(2):314–321.

24. Ivers N, Jamtvedt G, Flottorp S, et al. Audit and feedback: effects on professional practice and healthcare outcomes. Cochrane Database Syst Rev. Jun 13 2012;(6):Cd000259. doi:10.1002/14651858.CD000259.pub3

25. Pillai M, Posada J, Gardner RM, et al. Measuring quality-of-care in treatment of young children with attention-deficit/hyperactivity disorder using pre-trained language models. Journal of the American Medical Informatics Association. 2024;doi:10.1093/jamia/ocae001

26. Sarkar U, Bates DW. Using Artificial Intelligence to Improve Primary Care for Patients and Clinicians. JAMA Internal Medicine. 2024;doi:10.1001/jamainternmed.2023.7965

