## Supplementary material for "Leveraging a Large Language Model to Assess Quality-of-Care: Monitoring ADHD Medication Side Effects": eSupplement

**e-Supplement**

**Supplementary Figure 1.** Study cohort flowchart.

**e-methods supplement.**

**Supplementary Figure 2.** Classification of side effect inquiry using LLaMA

**Supplementary Figure 1**. Study cohort flowchart.

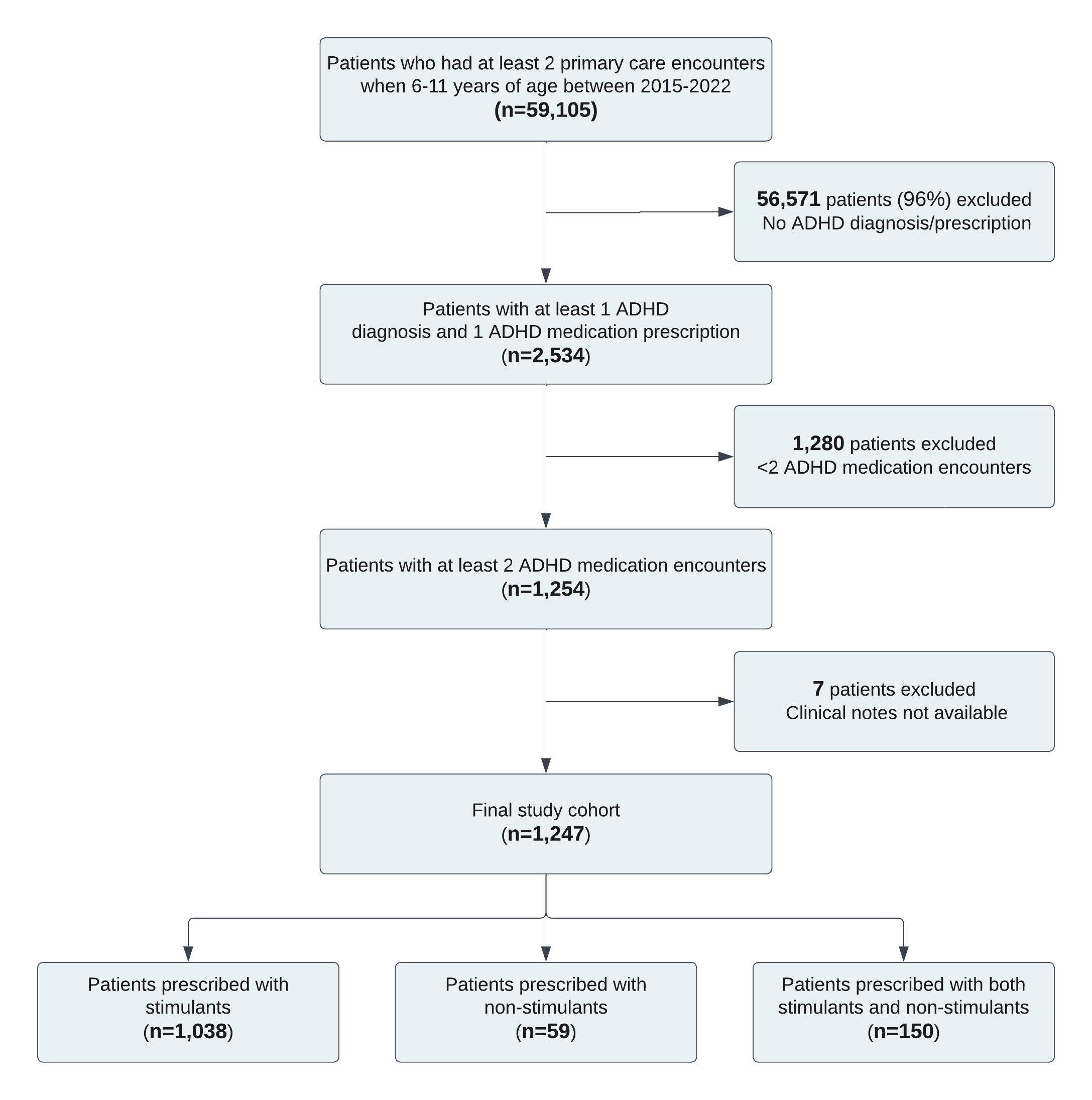

**e-methods supplement**

*ADHD-related encounters and medications*

ADHD-related encounters were identified by visit diagnoses that had the Observational Medical Outcomes Partnership Common Data Model (OMOP CDM) code for ADHD (concept id = 43409) or its descendant concepts, except for diagnoses of hyperkinesis (438132, 437261, and descendants) that were excluded. For each patient, a first ADHD diagnosis was defined as the first ADHD-related encounter in the study period (10/2015-12/2022).

ADHD medication prescription was identified by OMOP CDM code for methylphenidate (705944), guanfacine (1344965), dextroamphetamine (719311), dexmethylphenidate (731533), clonidine (1398937), atomoxetine (742185), amphetamine (714785), or any descendant concept.

*Data Preprocessing*

The data preprocessing steps included both the cleaning and sectioning of the clinical notes. Cleaning involved the elimination of punctuation, stop words, excessive whitespace, and digits. Subsequently, the notes underwent sectioning to adhere to the limitations imposed by the LLMs on note lengths. Specifically, notes were segmented into three distinct sections: subjective, objective, and assessment. Each section received distinct labels based on the regions highlighted by clinicians.

*Text Embeddings and Model Training*

Text embeddings were generated using the newly available LM Meta AI (LLaMA), followed by the addition of a dense layer for classifying clinical notes as either including medication side effect (SE) documentation or not (see **e-Figure**). The annotated notes were split into training (60%), hyperparameter optimization (20%), and testing (20%) sets, ensuring no overlap between them. All note sections from a single patient were allocated to only one of these subsets. Hyperparameter tuning was performed using grid search, with the following parameter ranges: learning rate: [0.001, 0.0001], batch size: [32, 64, 128], epochs: [20, 50], dense layer nodes: [64, 128, 256], and dropout rate: [0, 0.1, 0.2].

*Model Evaluation and Encounter- and Patient-level Predictions*

After obtaining section-level predictions, they were aggregated into note-level predictions by selecting the maximum prediction from each section. The performance of the note-level models was evaluated on the test set using various metrics, including sensitivity, specificity, and the area under the receiver operating characteristic curve (AUC). Model thresholds were selected to maximize sensitivity and minimize the false negative rate on the training set. Subsequently, encounter and patient-level predictions were derived. Each note was initially linked to the corresponding encounters. If SE was documented in at least one note, encounters were classified as 'SE inquiry’; otherwise, they were labeled as 'no SE inquiry.' Similarly, patients were classified as 'SE inquiry' if SE was documented in at least one encounter. Patients with no SE documentation in any encounter were labeled as 'no SE inquiry'. All experiments were conducted using Python with the PyTorch and Keras libraries.

**Supplementary Figure 2** Classification of side effect inquiry using LLaMA

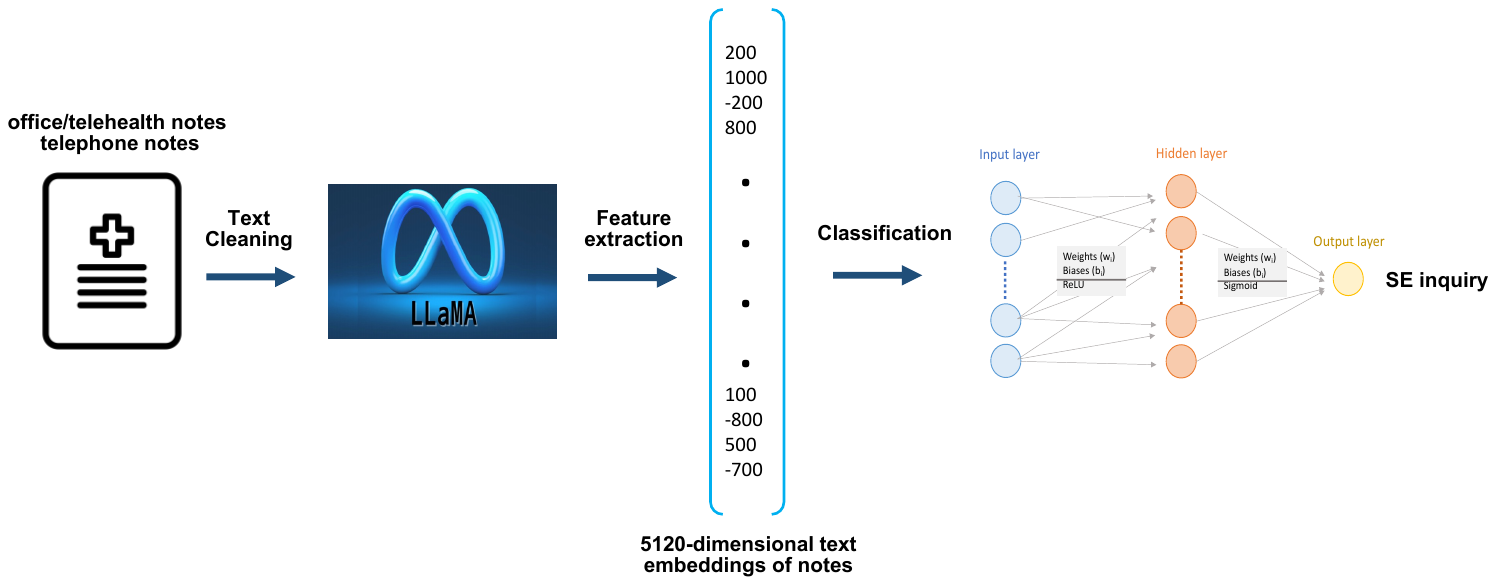
